# Quality of life and its determinants among individuals with type 2 diabetes mellitus in rural Bangladesh

**DOI:** 10.1101/2025.06.11.25329444

**Authors:** Lucie Sabin, Sanjit Kumar Shaha, Ali Kiadaliri, Abdul Kuddus, Carina King, Xingzuo Zhou, Naveed Ahmed, Hannah Jennings, Joanna Morrison, Kohenour Akter, Tasmin Nahar, Kishwar Azad, Edward Fottrell, Hassan Haghparast-Bidgoli

## Abstract

**Background:** Type 2 diabetes mellitus (T2DM) is a growing public health issue in Bangladesh, with the rural population facing significant barriers to diagnosis and care. While previous studies conducted in urban areas have examined the health-related quality of life (HRQoL) of people with diabetes, evidence from rural areas remains limited.

**Methods:** We analysed data from 1,574 adults with diabetes in rural Bangladesh, from the DMagic cluster randomised trial. HRQoL was assessed using the EQ-5D-3L instrument and the visual analogue scale. HRQoL was compared between people with a previous diagnosis of diabetes and those who were unaware of their condition prior to the study. Multivariable linear regressions and modified Poisson regressions were used to examine the associations between HRQoL and socio-demographic factors, risk behaviours, and comorbidities.

**Results:** Results indicate that socio-demographic, economic and health factors are associated with HRQoL. The main factors associated with higher HRQoL and clinically relevant were Higher levels of education, being male and belonging to a higher wealth tertile. Individuals with diabetes unaware of their condition were less likely to report problems in mobility, self-care, pain/discomfort and usual activities.

**Conclusion:** The findings emphasised the need for targeted interventions for high-risk groups, especially individuals with low socio-economic levels.

## Introduction

Type 2 Diabetes mellitus (T2DM) is a major global health issue leading to significant morbidity and premature mortality (1). The Southeast Asia region has one of the highest rates of T2DM globally, largely driven by India, Indonesia, and Bangladesh (2). In 2019, approximately 8.4 million adults in Bangladesh were reported to have T2DM (2) and projections suggest that this number could double by 2045 (2), with some estimates indicating that this could happen even sooner (3,4).

While clinical measures offer a useful estimate of disease control, the primary goal of health interventions is to enhance quality of life (5). Health-related quality of life (HRQoL) encompasses individuals’ physical and physiological states (6) and provides an understanding of the multifaceted impact of a disease on patients’ daily lives (7). People with T2DM experience a deterioration in HRQoL due to complications associated with the disease, prolonged use of medication and recommended lifestyle changes (8–10). Studies conducted in Southeast Asia have highlighted various determinants of HRQoL in patients with T2DM, with studies conducted in Bangladesh (5) and India (11) identifying diet, exercise and insulin use as key factors. Several studies conducted in Bangladesh (12–14), Nepal (9), Pakistan (15,16) and India (11,17–20) noted socioeconomic inequalities in HRQoL among T2DM patients, while other studies highlight the role of co-morbidities (8,9,14,17,18) and disease complications (11,13,20–23) in determining HRQoL among people with diabetes.

However, all assessments of HRQoL in Bangladesh among people with diabetes have focused primarily on urban populations, leaving a significant gap in understanding the experiences of people living in rural areas. Access to healthcare for the screening, diagnosis, and treatment of diabetes in rural Bangladesh is limited (24), and many patients with this condition remain undiagnosed (3), potentially influencing the impact of diabetes on their HRQoL. To address this gap, our study aimed to investigate factors associated with HRQoL of a rural population with diabetes in Bangladesh. We will also compare HRQoL between previously diagnosed people who potentially benefited from diabetes management and people with diabetes who were unaware of their diabetes status. As diabetes is a chronic disease, understanding the factors that influence the HRQoL of people with diabetes is essential to improving long-term disease management, tailoring healthcare interventions and improving the well-being of people with diabetes in rural areas.

## Methods

### Study design, setting and participants

The DMagic study was a three-arm cluster randomised controlled trial conducted in four rural sub-districts of Faridpur, Bangladesh. For our analysis, we used data from the DMagic endline evaluation survey, which included interviews with 11,454 permanent residents, males and non-pregnant females, aged 30+ years between Jan 16, and April 30, 2018. Using stratified randomisation, the 96 villages in Faridpur were divided equally into three groups: mHealth intervention, participatory learning and action (PLA) intervention, and control. In the mHealth group, participants received voice messages twice a week for 14 months. These messages aimed to improve health behaviours by raising awareness of diabetes, providing information about its symptoms, prevention and management, as well as practical advice to reduce the risk of developing diabetes and its complications (25). The PLA intervention consisted of monthly meetings conducted in a four-phase cycle. Participants identified and prioritised risk factors for T2DM, devised feasible strategies to address these risks, implemented the strategies with their communities and evaluated their effect. The findings from the DMagic trial showed that the PLA intervention was effective in preventing type 2 diabetes. Further details of the interventions and trial results are available in previous publications (25–27).

### Dependent variables

During the survey, data on quality of life was collected using the Bengla-translated version of the EQ-5D-3L questionnaire (28) among a sample of previously diagnosed people with diabetes and people with diabetes who were unaware of their diabetes status. Diabetes was defined as fasting blood glucose greater than or equal to 7 mmol/L, 2-hour postprandial blood glucose or oral glucose tolerance test (OGTT) greater than or equal to 11.1 mmol/L, or random blood glucose greater than or equal to 11.1 mmol/L in people clinically identified as diabetic. The EQ-5D-3L is a standardised instrument designed to measure self-reported HRQoL (29). It consists of a simple questionnaire that assesses five dimensions: mobility, self-care, usual activities, pain/discomfort, and anxiety/depression. Each dimension consists of three levels of severity: no problems, some problems, and extreme problems. We combined ‘some problems’ and ‘extreme problems’ given only a few extreme problems were reported. We determined the EQ-5D-3L index scores by weighting each dimension using the United Kingdom (UK) value set (30), selected to facilitate comparison with prior studies conducted in Bangladesh (12–14).

Data were also collected on self-rating of overall health status using a visual analogue scale (EQ-VAS) which allows participants to rate their health on a continuous scale ranging from 0 (representing the worst imaginable health state) to 100 (representing the best imaginable health state) (31). The VAS provides a subjective measure of an individual’s perceived health status.

### Independent variables

Based on a literature review of determinants of HRQoL in Southeast Asia, we defined independent variables a priori, including: sex, age, education (no formal education, primary, secondary, and tertiary), religion (Muslim, other) occupation (unemployed, housework/housewife, manual labour, non-manual labour), marital status (currently married or not), wealth tertiles (household-level based on a principal components analysis of asset ownership), risk factors (measured overweight and self-reported current use of tobacco), and having diabetes complications or co-morbidities (foot ulcers, hypertension, heart disease, kidney problems including frequent urination and ankle/leg swelling, blurred/loss of vision, ulcers, stroke and incontinence). Overweight was determined based on measured height and weight, using the WHO thresholds for South Asia, where overweight is defined as a body mass index (BMI) ≥23 kg/m².

### Statistical analysis

We employed descriptive and multivariable analyses to assess the associations between independent variables with the HRQoL (dependent variables). Firstly, we used multiple linear regression to investigate the association of independent variables with the EQ-VAS. Then, a similar analysis was conducted with the dependent variable being the EQ-5D-3L index scores, which the results from the latter analysis are presented in supplementary material 1. Group differences were interpreted using the minimal clinical important difference (MCID). For any differences smaller than the MCID, we considered them clinically irrelevant, even if statistically significant. Based on an average EQ-VAS of 73 and an average EQ-5D-3L index score of 0.71, we considered the MCIDs to be 4.8 [95% CI=3.1;6.6] and 0.10 [95% CI=0.07;0.12] respectively (32). Thirdly, we used modified Poisson regression to investigate the association of independent variables with the five dimensions of the EQ-5D-3L. These three models were applied separately to a sample of previously diagnosed people with diabetes and a sample of people with diabetes who were unaware of their diabetes status. We assessed the differences between the two samples using the Hausman test.

### Ethical approval

Ethical approval for D-Magic trial was obtained from University College London (4766/002) and the Diabetic Association of Bangladesh (BADAS-ERC/EC/t5100246). All survey participants provided written or thumb-print informed consent.

## Results

### Descriptive statistics

Descriptive statistics for the sample are presented in Table 1. The total sample size was 1,574 people with diabetes of whom 399 individuals had their diabetes diagnosed before the study and 1,175 individuals who met the definition for diabetes who were unaware of their diabetes status prior to our survey. The age distribution showed that the most participants were in the 40-49 age group, 30-39 age group, followed by the 50-59 age group, with frequencies decreasing in the older age groups. 64.8% of the sample were male. Most participants had no formal education, were housewives, married and Muslim. There was a low proportion of people currently using tobacco and 51% were overweight, with a higher proportion among those already diagnosed with diabetes. The most frequently reported coexisting conditions were gangrene and ulcers, while heart disease was the least common.

**Table 1.** Participant characteristics based on whether they were aware of their diabetes diagnosis.

| **Variables** | **Whole sample** |
| --- | --- |
| **Age** |  |
| 30-39 years | 366 (23.3%) |
| 40-49 years | 421 (26.7%) |
| 50-59 years | 352 (22.4%) |
| 60-69 years | 310 (19.7%) |
| 70 and more | 125 (7.9%) |
| **Sex** |  |
| Male | 554 (64.8%) |
| Female | 1,020<br>(35.2%) |
| **Education** |  |
| None | 685 (43.5%) |
| Primary | 748 (47.5%) |
| Secondary and more | 141 (9.0%) |
| **Marital status** |  |
| Married | 231 (14.7%) |
| Not married | 1,343<br>(85.3%) |
| **Religion** |  |
| Muslim | 187 (11.9%) |
| Other | 1,387<br>(88.1%) |
| **Occupation** |  |
| Not working | 177 (11.3%) |
| **Housework/housewife** | 934 (59.3%) |
| Manual labour | 312 (19.8%) |
| Non-manual labour | 151 (9.6%) |
| **Wealth tertiles** |  |
| Poorest | 572 (36.3%) |
| Middle | 481 (30.6%) |
| Least poor | 521 (33.1%) |
| **Current use of tobacco (self-reported)** |  |
| Yes | 188 (11.9%) |
| No | 1,386<br>(88.0%) |
| **Overweight (measured)** |  |
| Yes | 810 (48.5%) |
| No | 764 (51.5%) |
| **Eye problems** |  |
| Yes | 453 (28.8%) |
| No | 1,121<br>(71.2%) |
| **Kidney problems** |  |
| Yes | 285 (18.1%) |
| No | 1,289<br>(81.9%) |
| **Stroke** |  |
| Yes | 471 (29.9%) |
| No | 1,103<br>(70.1%) |
| **Incontinence** |  |
| Yes | 452 (28.7%) |
| No | 1,122<br>(71.3%) |
| **Gangrene** |  |
| Yes | 733 (46.6%) |
| No | 841 (53.4%) |
| **Ulcers** |  |
| Yes | 716 (45.5%) |
| No | 858 (54.5%) |
| **Hypertension** |  |
| Yes | 373 (23.7%) |
| No | 1,201<br>(76.3%) |
| **Heart disease** |  |
| Yes | 145 (9.2%) |
| No | 1,429<br>(90.8%) |
| **Total** | **1,574** |

Distributions of EQ-VAS and EQ-5D-3L index scores are presented in Figure 1 and Figure 2. Among the whole sample of individuals with diabetes, the mean EQ-VAS was 73 [95% CI=71.8;73.3]. The mean scores were 70 [95% CI=68.6;71.6] for people diagnosed with diabetes before the study versus 73 [95% CI=72.4;74.3] for those who were unaware of their diabetes at the time of study. The mean UK EQ-5D-3L index score was 0.69 [95% CI=0.66;0.72] for people aware of their diabetes status and 0.72 [95% CI=0.71;0.74] for those unaware. The mean UK EQ-5D-3L index score for the whole sample was 0.71 [95% CI=0.70;0.73].

**Figure 1.**
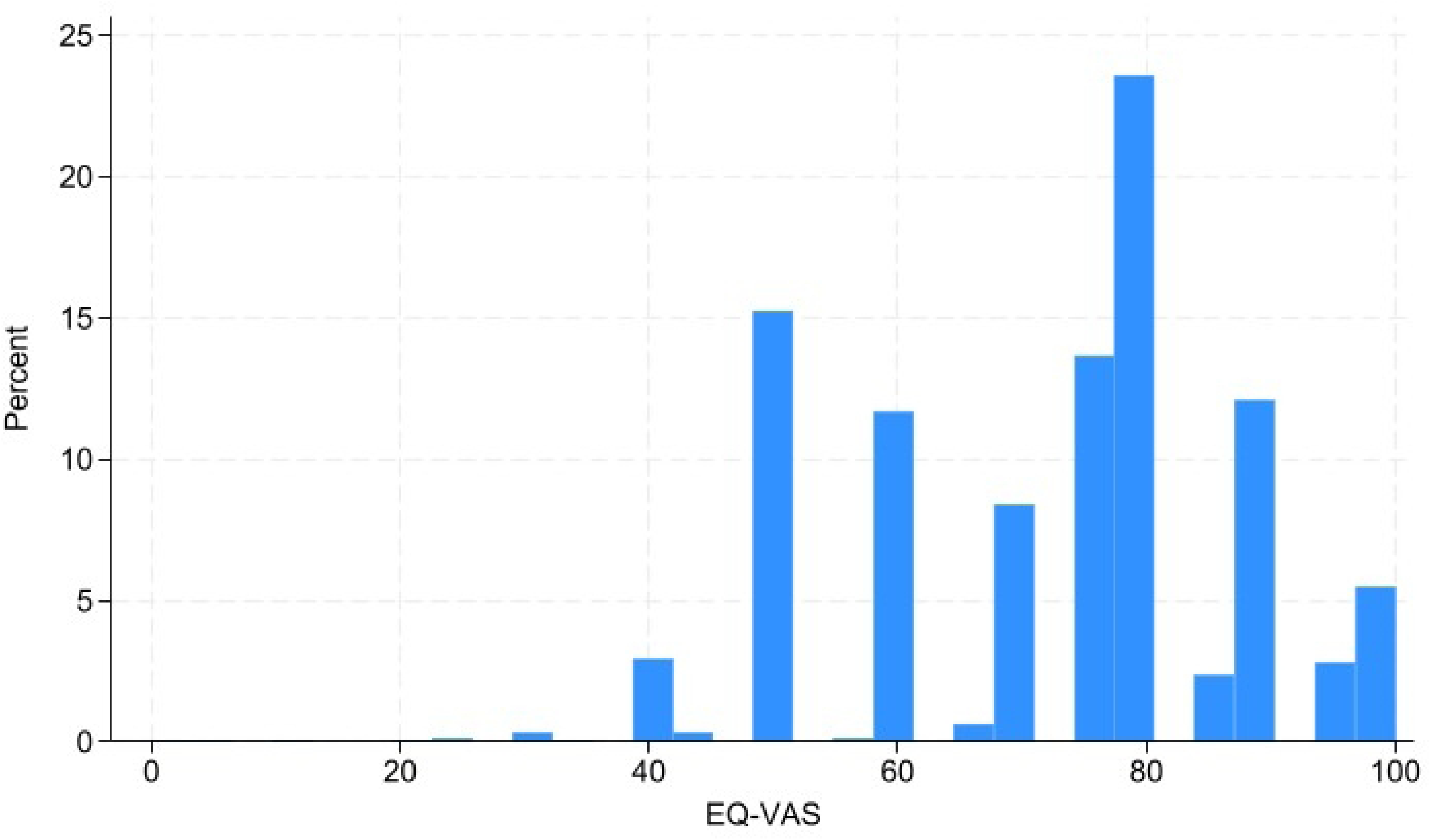
Distribution of EQ-VAS in percentage.

**Figure 2.**
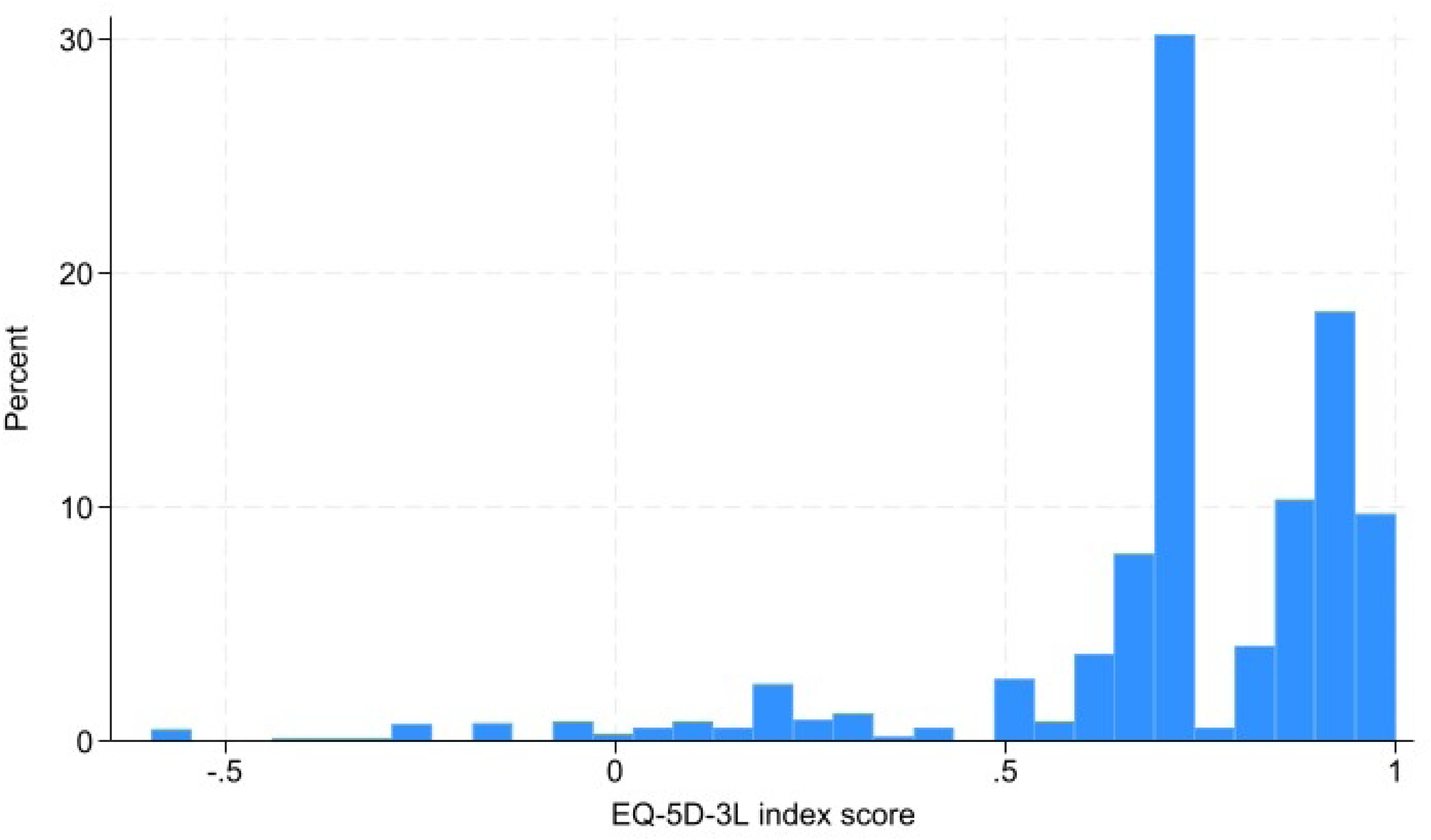
Distribution of EQ-5D-3L index scores in percentage.

Table 2 presents the distribution of the responses to the EQ-5D-3L. Pain/discomfort (60.2%) followed by anxiety/depression (58.4%) were the most affected dimensions, while self-care (8.2%) was the least affected.

**Table 2.** Distribution of responses in people with T2DM according to the severity of problems across the five dimensions of the HRQoL.

| Variables | Whole sample | People unaware of<br>their diabetes status | People aware of<br>their diabetes status |
| --- | --- | --- | --- |
| **Mobility** |  |  |  |
| No problem | 1,309 (83.2%) | 1,012 (86.1%) | 297 (74.4%) |
| Some/extreme problems | 265 (16.8%) | 163 (13.9%) | 102 (25.6%) |
| **Self-care** |  |  |  |
| No problem | 1,445 (91.8%) | 1,087 (92.5%) | 358 (89.7%) |
| Some/extreme problems | 129 (8.2%) | 88 (7.5%) | 41 (10.3%) |
| **Usual activities** |  |  |  |
| No problem | 1,235 (78.5%) | 955 (81.3%) | 280 (70.2%) |
| Some/extreme problems | 339 (21.5%) | 220 (18.7%) | 119 (29.8%) |
| **Pain and discomfort** |  |  |  |
| No problem | 626 (39.8%) | 492 (41.9%) | 134 (33.6%) |
| Some/extreme problems | 948 (60.2%) | 683 (58.1%) | 265 (66.4%) |
| **Anxiety and depression** |  |  |  |
| No problem | 622 (41.6%) | 421 (36.9%) | 201 (56.5%) |
| Some/extreme problems | 875 (58.4%) | 720 (63.1%) | 155 (43.5%) |

### Key findings

Variables included in the model examining the association with the EQ-VAS are presented in Table 3, while those in the model analysing the association with the EQ-5D-3L dimensions are detailed in Supplementary Material 1. Age, currently using tobacco and having comorbidities were slightly negatively associated with the EQ-VAS. B being Muslim was also negatively associated with the EQ-VAS. Higher education levels were positively associated with the EQ-VAS. Similarly, being a male, being currently married and belonging to a higher wealth tertile had a positive association with EQ-VAS. Working, whether as a manual worker or as a professional/business worker or being a housewife was positively associated with EQ-VAS. Among people with diabetes, being unaware of their condition was positively associated with EQ-VAS.

**Table 3.** Associations between sociodemographic and co-morbidities and EQ-VAS among people with.

|  | **Coef.** | **95% CI** |
| --- | --- | --- |
| Age | -0.13 | -0.17,-0.08 |
| Male | 4.7 | 0.97,8.4 |
| Education level |  |  |
| None | REF. |  |
| Primary | 2.98 | 1.28,4.68 |
| Secondary & higher | 5.80 | 2.63,8.96 |
| Currently married | 2.03 | -0.34,4.40 |
| Muslim | -1.27 | -3.63,1.08 |
| Occupation |  |  |
| Not working | REF. |  |
| Housewife/housework | 10.32 | 6.87,13.77 |
| Manual | 8.81 | 5.64,11.98 |
| Professional/business | 7.28 | 3.65,10.91 |
| Wealth tertiles |  |  |
| Poorest | REF. |  |
| Middle | 3.32 | 1.46,5.18 |
| Least poor | 3.99 | 2.00,5.98 |
| Current user of tobacco | -0.47 | -3.20,2.25 |
| Comorbidities | -0.34 | -0.79,-0.11 |
| Unaware of their diabetes status | 3.70 | 1.93,5.47 |
| Constant | 62.54 | 56.91,68.17 |

Notes: REF.= reference category. Coef.= coefficients. CI= confidence interval.

Factors associated with reporting problems across the five dimensions of the EQ-5D-3L are reported in Table 4. Men were less likely than women to report some/extreme problems among all dimensions except anxiety/depression and self-care. People with higher education, people working as well as currently married were less likely to report problems across the five dimensions than their counterparts. Similarly, individuals with diabetes from middle and least-poor wealth tertiles were less likely to report problems across the five dimensions than people belonging to the poorest tertile. Muslim people were more likely to report problems in all dimensions except self-care. Currently using tobacco demonstrated mixed effects depending on the dimension with people currently using tobacco being more likely to report problems with anxiety/depression than individuals who do not use tobacco. In contrast, individuals who currently use tobacco were less likely to report problems regarding mobility, self-care and pain/discomfort. Individuals with diabetes unaware of their status were less likely to report problems in all dimensions except anxiety/depression than people aware of their diabetes status. An increasing number of comorbidities was associated with a higher probability of reporting problems with usual activities and pain/discomfort. In contrast, an increasing number of comorbidities was associated with a lower probability of reporting problems with self-care and anxiety/depression.

**Table 4.** Factors associated with reporting some/extreme problems for each dimension of the EQ-5D-3L among people diagnosed with diabetes (N=1,574)

| Independent variables | Mobility |  | Self-care |  | Usual activities |  | Pain and discomfort |  | Anxiety and depression |  |
| --- | --- | --- | --- | --- | --- | --- | --- | --- | --- | --- |
|  | Coef. | 95% CI | Coef. | 95% CI | Coef. | 95% CI | Coef. | 95% CI | Coef. | 95% CI |
| Age | 1.02 | 1.01,1.03 | 1.02 | 1.00,1.04 | 1.02 | 1.00,1.03 | 1.00 | 1.00,1.01 | 1.00 | 1.00,1.00 |
| Male | 0.79 | 0.56,1.13 | 1.20 | 0.69,2.07 | 0.91 | 0.68,1.22 | 0.81 | 0.67,0.96 | 1.21 | 0.99,1.46 |
| Education level |  |  |  |  |  |  |  |  |  |  |
| None | REF. |  |  |  |  |  |  |  |  |  |
| Primary | 0.71 | 0.55,0.92 | 0.72 | 0.49,1.06 | 0.64 | 0.52,0.79 | 0.88 | 0.81,0.97 | 0.87 | 0.80,0.95 |
| Secondary & higher | 0.48 | 0.29,0.81 | 0.57 | 0.26,1.25 | 0.36 | 0.21,0.61 | 0.65 | 0.52,0.82 | 0.68 | 0.54,0.87 |
| Currently married | 0.85 | 0.63,1.14 | 0.57 | 0.36,0.90 | 0.85 | 0.66,1.09 | 1.00 | 0.90,1.10 | 0.91 | 0.80,1.02 |
| Muslim | 1.32 | 0.92,1.87 | 0.88 | 0.55,1.41 | 1.00 | 0.78,1.28 | 1.09 | 0.96,1.24 | 1.09 | 0.94,1.26 |
| Occupation |  |  |  |  |  |  |  |  |  |  |
| Not working | REF. |  |  |  |  |  |  |  |  |  |
| Housewife/housework | 0.38 | 0.27,0.53 | 0.20 | 0.11,0.36 | 0.40 | 0.31,0.52 | 0.82 | 0.72,0.93 | 1.00 | 0.84,1.19 |
| Manual | 0.28 | 0.17,0.47 | 0.16 | 0.11,0.52 | 0.34 | 0.23,0.50 | 0.64 | 0.53,0.77 | 0.85 | 0.73,0.99 |
| Professional/business | 0.40 | 0.23,0.74 | 0.19 | 0.11,0.80 | 0.31 | 0.18,0.54 | 0.75 | 0.72,0.93 | 0.93 | 0.75,1.14 |
| Wealth tertiles |  |  |  |  |  |  |  |  |  |  |
| Poorest | REF. |  |  |  |  |  |  |  |  |  |
| Middle | 0.91 | 0.63,1.31 | 0.91 | 0.65,1.37 | 0.89 | 0.72,1.09 | 0.90 | 0.82,0.99 | 0.92 | 0.84,1.00 |
| Least poor | 0.73 | 0.49,1.08 | 0.73 | 0.54,1.24 | 0.81 | 0.65,1.01 | 0.91 | 0.82,1.00 | 0.83 | 0.74,0.93 |
| Current user of tobacco | 0.67 | 0.32,1.37 | 0.67 | 0.32,1.44 | 1.00 | 0.70,1.41 | 0.96 | 0.79,1.15 | 1.04 | 0.90,1.19 |
| Comorbidities | 1.03 | 0.98,1.09 | 0.95 | 0.87,1.04 | 1.02 | 0.97,1.08 | 1.03 | 1.00,1.05 | 0.88 | 0.86,0.90 |
| Unaware of their diabetes status | 0.72 | 0.51,1.00 | 0.72 | 0.53,1.06 | 0.60 | 0.50,0.72 | 0.86 | 0.79,0.93 | 1.24 | 1.09,1.40 |

*Notes: REF. stands for the reference category. Coef.= coefficients that should be interpreted as prevalence rate ratio. CI= confidence* *interval.*

## Discussion

In the current study, we evaluated factors associated with the HRQoL among individuals with diabetes in rural Bangladesh using the EQ-5D-3L and EQ-VAS instruments. We found associations between socio-demographic, economic, and health factors and the HRQoL. Higher levels of education, being male, married, belonging to a higher wealth tertile and working were positively associated with HRQoL, while age, smoking and co-morbidities were negatively associated.

We found that the mean EQ-VAS among people with diabetes was 73 out of 100, reflecting a relatively high level of HRQoL compared to previous studies conducted in Bangladesh (13,14). Among a sample of people with diabetes, Safita (2016) (13) found an unadjusted EQ-VAS mean of 69.0. We also found a higher level of HRQoL when expressed in terms of mean UK EQ-5D-3L (0.71) compared to previous studies conducted in Bangladesh. Saleh (2015) (12) observed a mean UK EQ-5D index score of 0.55.

Our results align with other studies reporting lower HRQoL in urban compared to rural areas among people with diabetes (12,33). The higher scores in our study could reflect differences in settings, as our research was conducted in rural areas where community support might be stronger (12,34). Additionally, differences in the results with other previous studies may be due to HRQoL being a variable that fluctuates over time, potentially influenced by factors like disease progression, access to healthcare, and changes in personal or socioeconomic situations (35).

Among the EQ-5D-3L dimensions, we found that pain/discomfort (60.2%), along with anxiety/depression (58.4%), were the most frequently reported problems among individuals with diabetes, highlighting the significant impact of the disease on both physical and mental health. Although the percentages were lower in our study, this is consistent with Saleh (2015) (12), which found that pain/discomfort and anxiety/depression were among the categories most affected, with 72.8% and 73.6% of individuals with diabetes reporting some/extreme problems, respectively. Similarly, Barua (2021) (14) reported that, among the five dimensions assessed, the highest rates of problems were 79.8% for anxiety/depression and 77.7% for pain/discomfort.

Among people with diabetes, being unaware of their condition was positively associated with EQ-VAS. Although these results aligned with findings from previous studies that highlight the impact of chronic diseases such as diabetes on quality of life (36,37), they were not clinically significant (group differences were smaller than the MCID). We found that individuals with diabetes unaware of their condition were clinically significantly less likely to report problems in mobility, self-care, pain/discomfort and usual activities. This may suggest that people who are unaware that they have diabetes experience fewer physical or psychological constraints linked to the management of their disease. These associations could also be linked to lifestyle changes, medication compliance and the ongoing self-management required to control diabetes, as well as awareness of potential long-term complications (38). In addition, a diagnosis may introduce financial stress due to the costs of medical consultations, medications, and necessary lifestyle adjustments, which could further contribute to the reported differences in well-being. These results emphasise the importance of timely and supportive interventions to help newly diagnosed patients manage the emotional and physical challenges of diabetes. Moreover, people who are unaware that they have diabetes have higher levels of anxiety/depression. They may suffer psychological distress, potentially due to underlying health problems or the uncertainty of an undiagnosed illness.

Several socio-demographic factors were found to be associated with HRQoL outcomes. Men reported higher HRQoL compared to women and this was clinically significant. These findings align with other studies conducted in Bangladesh (12–14). This disparity can be attributed to differences in men’s and women’s perceptions of health, barriers to access healthcare for women, and differences in social roles (39). Women are often faced with greater caregiving responsibilities and greater financial dependence, which can limit their access to healthcare services (40). In addition, societal expectations may limit women’s ability to seek care or prioritise their own health, as they may feel obliged to prioritise family or household needs, thus influencing their self-reported HRQoL outcomes.

In line with another study previously conducted in Bangladesh (12), we found that age was slightly negatively associated with HRQoL. This finding was not clinically significant. This trend can be attributed to the age-related decline in physical functioning, which naturally limits mobility and daily activities and increases susceptibility to chronic pain (41). In addition, according to a global study, older people often suffer from a greater number of co-morbidities, which exacerbate physical limitations and discomfort (42). These findings highlight the importance of age-specific support strategies in diabetes management to help older people maintain their HRQoL.

Similarly to other studies conducted in South Asia, although not clinically significant, we found that being currently married was positively associated with higher HRQoL (15,17,19). The positive association of marriage with HRQoL can be explained by the emotional, financial and social support provided by spouses. In Asia, marriage can also bring wider family support and social legitimacy (43). Marriage often facilitates better adherence to treatment, as partners may encourage healthier behaviours and provide caregiving support (44), leading to better health outcomes and higher HRQoL. In line with these findings, we found that married individuals were less likely to report problems regarding self-care.

We found that higher education, paid or unpaid work and higher wealth were positively associated with HRQoL and this finding was clinically relevant. These findings were in line with previous studies conducted in South Asia (11,15,18,45) and Bangladesh (12,13). Considering clinical certainty, we also found that people with higher education and people with paid or unpaid work were less likely to report problems across the five dimensions than their counterparts. Care-seeking is likely to be better among those who are financially secure - in employment - than others. Education may enable a good standard of self-care and understanding of health information, ultimately contributing to better perceived health (46). This aligns with our findings that participants diagnosed before the study with secondary or higher education had a significantly lower probability of reporting problems with anxiety and depression than participants with no education. Similarly, the positive association of wealth/income with HRQoL may be explained by the fact that higher income allows better access to healthcare services, medications, and a healthier lifestyle (47). Individuals with higher incomes are also less likely to experience the financial strain that can accompany chronic disease, reducing stress and anxiety and thus improving overall quality of life (48). This study showed that participants with higher income were less likely to report problems with self-care, usual activities and anxiety/depression than people from the poorest households.

We found that more number of comorbidities was negatively associated with the HRQoL, even though this was not clinically significant. The negative impact of comorbidities on HRQoL was found in other studies in South Asia (8,9,14).

Our study has several limitations that should be acknowledged. Firstly, the use of self-reported data may introduce reporting bias, particularly for sensitive information. Specifically, the self-reporting of comorbidities may potentially result in misclassification during data collection. Secondly, the D-Magic trial did not collect data on the duration and severity of comorbidities. Third, the EQ-5D-3L, while widely used, may lack sensitivity to detect more nuanced changes in quality of life, particularly in populations with a high burden of co-morbidities. Moreover, our analysis was limited by the lack of data on several factors potentially influencing HRQoL, such as mental health status and access to healthcare. The use of the UK value set for the EQ-5D-3L may not accurately reflect the preferences of the Bangladeshi population, further complicating the interpretation of the results. Finally, using linear regression for EQ-5D-3L scores may not be the most appropriate method, as the distribution of the data may run counter to the assumptions of linear regression, which could lead to biased estimates.

These limitations aside, our findings contributed to the growing research area, exploring determinants of HRQoL in low- and middle-income countries. The large sample size and the use of validated, standardised tools to measure HRQoL allowed for robust investigations of factors influencing HRQoL among people with diabetes.

## Conclusion

This study investigated factors associated with HRQoL of people with diabetes in rural Bangladesh. Our results indicate that socio-demographic, economic and health factors are associated with HRQoL. Higher levels of education, being a male and belonging to a higher wealth tertile were positively and clinically significantly associated with HRQoL. Individuals with diabetes unaware of their condition were less likely to report problems in mobility, self-care, pain/discomfort and usual activities. Our findings emphasised the need for targeted interventions for high-risk groups, especially older individuals and those with multiple morbidities. The higher HRQoL in the current study conducted in rural areas compared with previous studies conducted in urban Bangladesh suggests that family support may play a role in alleviating some of the burdens associated with diabetes, a factor that could be exploited to improve diabetes management programmes in similar settings.

## Ethics approval

Ethical approval for D-Magic trial was obtained from University College London (4766/002) and the Diabetic Association of Bangladesh (BADAS-ERC/EC/t5100246). All survey participants provided written or thumb-printed informed consent.

## Availability of data and material

Data available upon request.

## Acknowledgement

The authors have no acknowledgements to declare.

